# Genomic Characterization of the *Comamonas kerstersii* Isolated from Diarrheal Patients in Bangladesh

**DOI:** 10.1101/2024.07.02.24309841

**Authors:** Noshin Ibnat Rib, Fariza Shams, Fahad Khan, Senzuti Sharmin, Sakib Abrar Hossain, Abdus Sadique, Jahidul Alam, Pronoy Debnath, Arman Hossain, Aura Rahman, Syeda Naushin Tabassum, Tahrima Saiha Huq, Maqsud Hossain

## Abstract

This study marks the first identification and genomic characterization of *Comamonas kerstersii* isolates from diarrheal patients in Bangladesh. C. kerstersii is a gram-negative opportunistic bacterium which has also been reported in several clinical samples, including urine, blood, stool, peritoneal liquid, sputum. We carried out the whole genome sequencing of three C. kerstersii isolates for the first time in Bangladesh with an aim to analyze genomic features in 3 different strains isolated from Bangladesh. Seven AMR genes, aph(6)-Id, aph(3’’)-Ib, mph(E), mph(F), msr(E), sul2, and tet(A), have been observed in all the strains. We identified several virulence factors including copper homeostasis genes, type IV pili and type VI secretion system. phage regions and IncQ1 plasmid were also detected.

## Introduction

The increasing number of persistent infections caused by relatively unknown non-fermenting gram-negative rods is a crucial source of concern in healthcare environments [1]. These emerging pathogens, which include species from the *Comamonas* genus, are opportunistic and pose severe risk to global health [2]. One of the little-known species in the *Comamonas* genus is *Comamonas kerstersii* [3]. *C. kerstersii* was first recognized as a distinct species in 2003, alongside *Comamonas aquatica* and *Comamonas terrigena* [3–6]. This species is a part of the rRNA homology group III and defined by the presence of a unique 23S rRNA gene and a distinct 16S rRNA gene sequence [4,7,8]. *C. kerstersii* was initially included in the *C. terrigena* complex [9]. Willems, et al.[10] studied the taxonomy of the genus *Comamonas* and suggested that *C. terrigena* contains 3 diiferent DNA group. Based on this information, Wauters, et al. (2003) studied the species and suggested that it comprise three different species based on 16S rRNA sequencing and DNA-DNA hybridization. They renamed DNA group II of *C. terrigena* as *C. aquatica* and DNA group III as *C. kerstersii*.

*C. kerstersi* species are non-fermenting, gram-negative, rod shaped bacillus [3,6,12–22]. 24 hours of incubation in ambient aerobic environment shows the growth of the species [13]. Better observation can be done if incubated for 48 hours and usually colonies have a diameter of 1-2 mm and seem translucent, smooth-surfaced, and nonpigmented [13,23]. *C. kerstersii* has previously been considered nonpathogenic (Jiang et al., 2018; Liu et al., 2020b; Opota et al., 2014b; Rong et al., 2022; Voitenkova et al., 2018). It is extensively found in soil, plants, and water with the ability to withstand aquatic environment [3,18,25,26]. But since 2013, the species have been found in several clinical samples like abdominal fluid, peritoneal fluid, blood, urine, stool and recently in sputum [6,9,14,15,18,23,25–28]. Several cases of intra-abdominal including appendicitis, peritonitis, diverticulosis, and few extra abdominal cases including urinary tract infection, bacteremia and pneumonia have been reported in number of papers [2,18,26]. In most cases the infections associated with *C. kerstersii* were polymicrobial [2,18,29]

Jiang et al [24] studied *C. kerstersii* 8943 and concluded in their study that the genome of *C. kerstersii* is circular with chromosome measuring 3,547,915 base pairs. The genome size is notably smallest among the *Comamonas* species studied aligning with previous findings. *C. kerstersii* 8943 has been identified to have numerous resistance genes, including tetA, strB, sul1, blaOXA-1, strA, sul2, catB3, and floR [24]. Another study shows that *C. kerstersii* is resistance to Ciprofloxacin [27]. The strain was also resistant to penicillin and ampicillin, and it tested positive for the *strA* and *strB* genes, which confer streptomycin resistance [24] while another study reported resistance to ampicillin [30]. Genetic analysis of the strain *C. kerstersii* 3132976 revealed three β-lactamase genes (CDSs H8N02_05890, H8N02_08740, and H8N02_17110) and among them H8N02_08740 and H8N02_17110 belong to class A and C β-lactamases, respectively [31]. In a recent paper, it reported that the *C. kerstersii* strain 121606 was resistant to almost all of the β-lactam class of antibiotic indicating that it has a extensive drug resistance phenotype[26].

Species of the genus *Comamonas* are associated with certain human infection and has diversified virulence factors [25,32]. To date, Wu, et.al. [17] one of the pioneers who studied virulence of *C. kerstersii* along with other species of the genus. The researchers discovered that *C. aquatica, C. terrigena*, and *C. kerstersii* constitute a unique cluster with common virulence features, such as genes for bacterial motility, adhesion, and chemotaxis signaling [17]. They have adhesin biosynthesis genes like IlpA and Hsp60, as well as biofilm-forming components, hemolysin genes and share stress proteins like as *ClpEP* and *SodB* for cellular defense [17].

Given its widespread distribution and increasing detection in clinical samples, it is critical to investigate the genomic characteristics and pathogenic potential of *C. kerstersii*, especially in regions with limited data. This study aims to bridge that knowledge gap by presenting the first comprehensive genomic analysis of *C. kerstersii* isolates from diarrheal patients in Bangladesh. It also provides new insights into the antimicrobial resistance and potential virulence factors of these isolates offering new insights into their antimicrobial resistance and potential virulence factors from Bangladesh.

## Material and Methods

### Sample Collection and Isolation

From March to November 2020, 17 stool samples were collected from the Dhaka Central International Medical College & Hospital (DCIMCH) to isolate *Vibrio cholerae.* The samples were carried in 5ml Cary Blair transport medium at 4°C and then incubated at 37° in 3ml Alkaline peptone water (APW) for enrichment. From the enriched culture, tellurite taurocholate gelatin agar (TTGA) and thiosulfate citrate bile-salts sucrose (TCBS) agar plates were inoculated and incubated overnight. Yellow colonies from TCBS and black-centred colonies from TTGA, presumptively identified by their morphology, were further streaked on gelatin agar (GA). Subsequent Gram staining and routine biochemical tests were performed for further characterization.

### Genome Assembly and Annotation

Isolates were inoculated in LB broth and grown at 37°C overnight for DNA extraction. Genomic DNA was extracted using QIAamp® DNA Mini Kit (QIAGEN, Hilden, Germany) according to manufacturer’s protocol. Library preparation and sequencing of 3 selected strains were carried out at Genome Research Institute of North South University (NGRI), Bangladesh. High molecular weight genomic DNA of the strains was used to prepare libraries using a Nextera DNA Library Prep Kit and employed for 250-bp paired-end whole genome sequencing with Illumina® MiSeq platform following the instruction provided by the manufacturer. High-quality paired-end Illumina sequencing data (Q ≥ 30) were obtained for the bioinformatic analysis. Quality control was performed on the sequencing data to ensure the integrity and accuracy of the information. SPAdes genome assembly software (version 3.15.5) [33] was used for the *de novo* assembly of the genome with particular filters employed to lower the amount of mismatches and short indels, improving the quality of the generated contigs. To decipher the genetic content and functional elements encoded within the assembled contigs, the PROKKA pipeline (version 45) [34]was employed for annotation. Rapid Annotation using Subsystem Technology (RAST) [35], an automated service for annotating genomes of bacteria and archaea, was also employed for annotation.

### Genomic Feature determination

Nine genomes (Supplementary Table 2) of *C. kerstersii* were obtained from the National Center for Biotechnology Information (NCBI). The obtained genomes were annotated using the Prokka pipeline [34]. The core genome alignment was generated using roary [36] and then used in FastTree [37] to construct a midpoint rooting phylogenetic tree and visualized using FigTree. Assembled genomes were visualized using the BLAST ring image generator (BRIG) [38]. Antimicrobial resistance genes were identified utilizing the Center for Genomic Epidemiology (CGE) based tool ResFinder (version 4.1) [39]. In this study, Virulence Factor Database (VFDB) (http://www.mgc.ac.cn/VFs/) has been used to identify and analyze virulence factors associated with the studied strains [40]. Identification of plasmids were conducted employing PlasmidFinder [41] and phage regions using PHASTEST [42] and Phigaro [43] respectively.

## Result

### Bacterial Identification

Of the 17 presumptive isolates, 14 demonstrated gelatin hydrolysis when tested on gelatin agar (GA) plates. The remaining three isolates did not exhibit gelatin hydrolysis, suggesting a different enzymatic profile. These three strains (NG13, NG14, and NG17) also produced a biochemical profile consistent with *Comamonas* species rather than *Vibrio cholera* (Table 1).

**Table 1.**
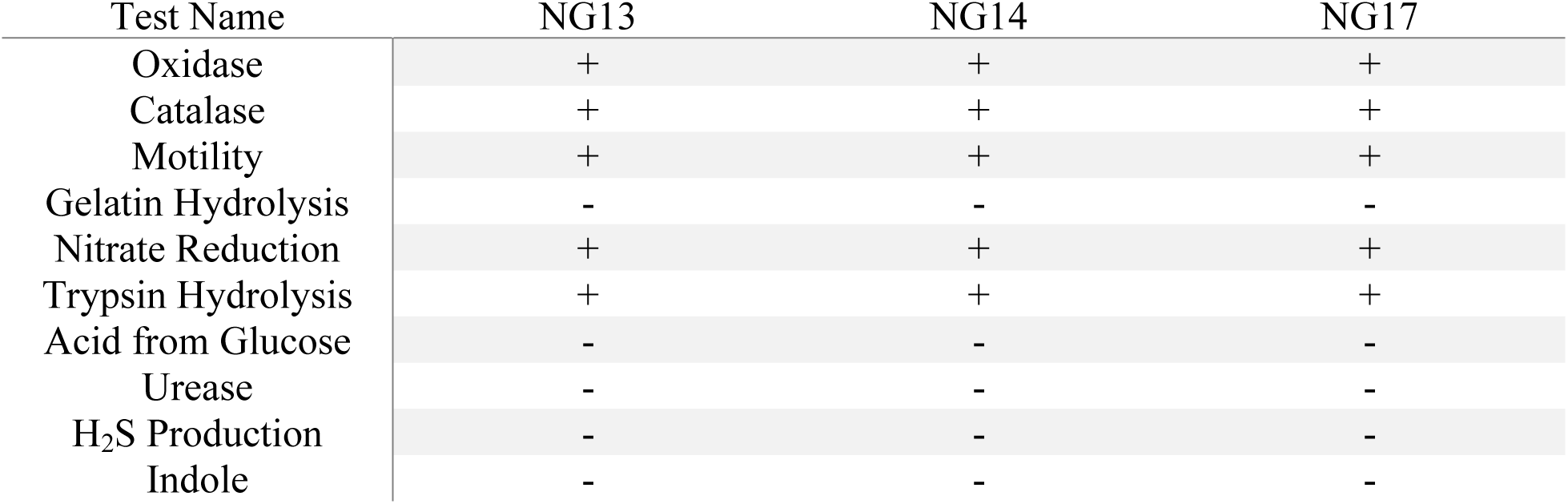
Biochemical test results of the three *C. kerstersii* isolates.

### Genomic Features

We conducted next-generation sequencing with the genomic DNA isolated from the three putative *Comamonas* strains NG13, NG14, and NG17. The draft genomes of strains NG13, NG14 and NG17 resulted in 50, 68, 81 contigs, respectively. The size of the assembled genomes of the 3 strains were 3.4 Mbp with approximately 59% GC content. NG13 and NG17 possess 94 tRNAs, NG14 possesses 97 tRNAs. All three strains contain 7 rRNAs and 1 tmRNA. The genomic features of the strains are summarized in Table 2.

**Table 2.** Genome assembly and annotation statistics of the three *C. kerstersii* isolates.

|  | NG13 | NG14 | NG17 |
| --- | --- | --- | --- |
| Genome Size (appx) | 34, 806, 663 | 34, 90, 070 | 34, 93, 044 |
| No. of contigs | 50 | 68 | 81 |
| GC (%) | 59.60 | 59.60 | 59.59 |
| No. of CDS | 3211 | 3215 | 3213 |
| No. of tRNA | 94 | 97 | 94 |
| No. of rRNA | 7 | 7 | 7 |
| No. of tmRNA | 1 | 1 | 1 |
| N50 (bp) | 181861 | 284992 | 221534 |

### Phylogeny Analysis

A core genome-based phylogenetic tree consisting of 12 *C. kerstersii* strains was constructed, which is shown in Figure 2. The tree shows two distinct clusters – Cluster A and Cluster B.

**Fig. 1.**
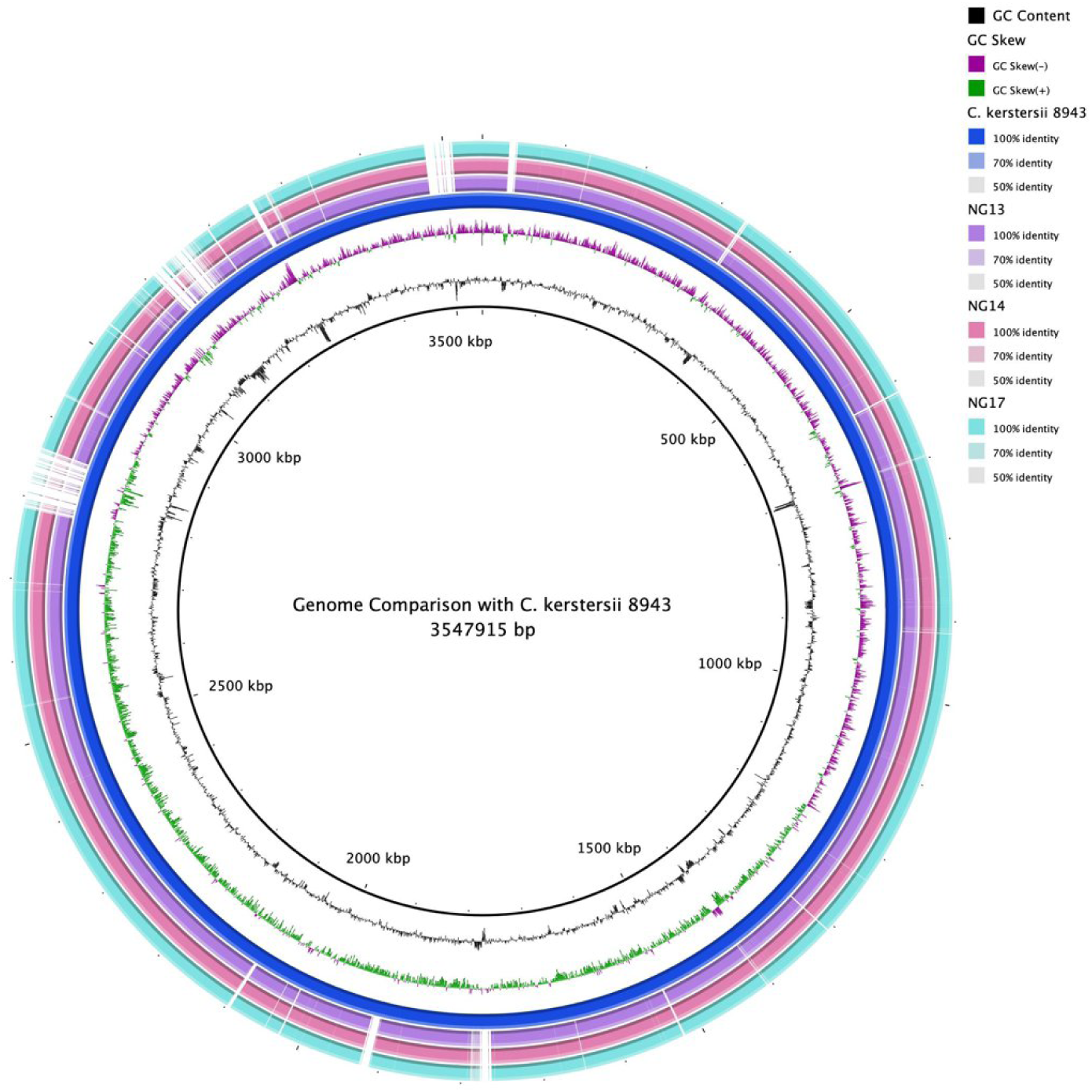
BRIG-obtained circular genomic maps of *C. kerstersii* strains. The homology rate is indicated by color saturation; blanks indicate no resemblance. The blue circle represents the reference genome *C. kerstersii* 8943.

**Fig. 2.**
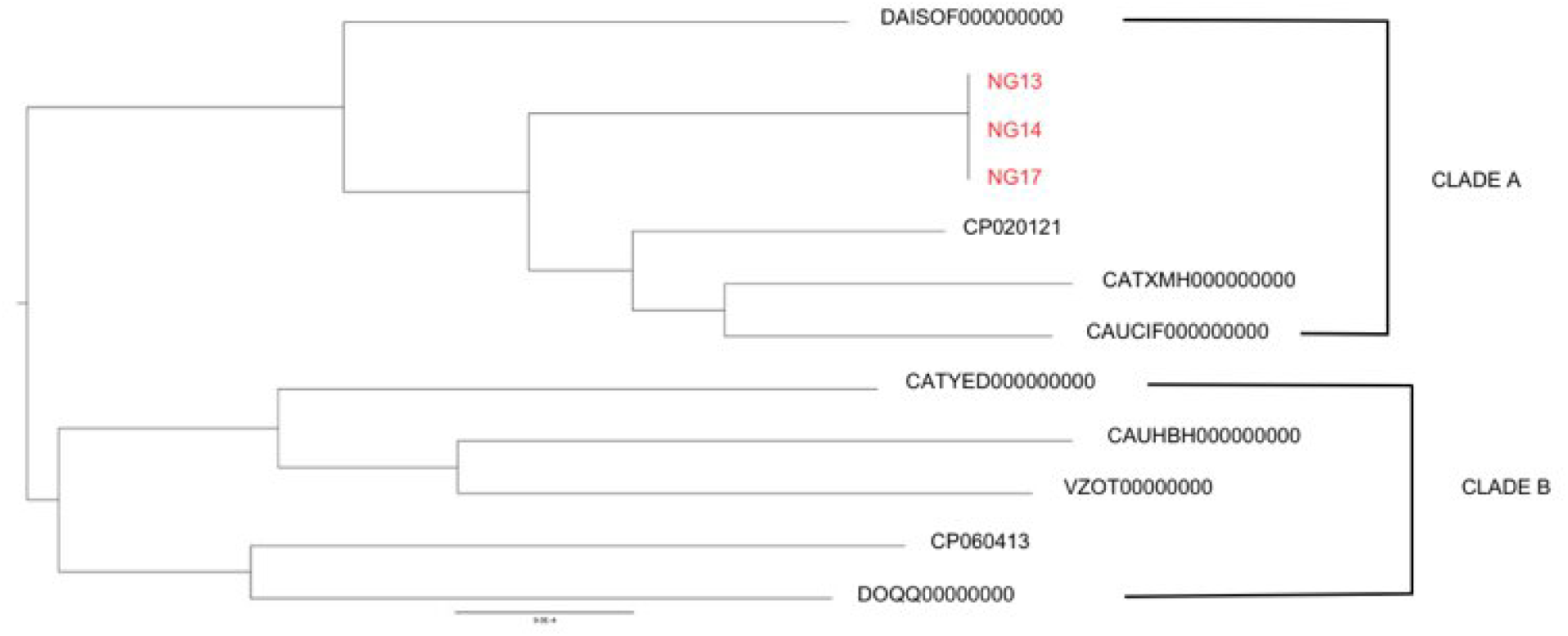
Mid-point rooted phylogenetic tree of publicly available *C. kerstersii* strains and sequenced isolates from Bnagladesh based on their core genome alignment. NG13, NG14 and NG17 belonging to CLADE A clustering together indicates their clonal character.

According to the phylogenetic tree, three *C. kerstersii* strains under study – NG13, NG14 and NG17, shared the same ancestor and belonged to Cluster A, with CP020121 being the closest relative. CP020121, also named as *C. kerstersii* 8943, was isolated from the effluent of peritoneal dialysis [24], highlighting the close relationship among human-originated isolates. All these strains share features like ampicillin resistance, the presence of sul2 and tetA genes conferring resistance to sulfonamide and tetracycline respectively. sulfonamides(Jiang et al. 2018).

The CP060413 strain, from the other cluster-Cluster B, was isolated from rectal swab [31] like the strains of the study. Even being from the same clinical source, The CP060413 branched distantly. This strain did not have any resistance genes other than 2 class A and 1 class C betalactamase genes.

The *C. kerstersii* strains used in the phylogenetic analysis were isolated either from human feces or as members of the gut microbiota. The presence of various resistance genes across different strains underscores the role of horizontal gene transfer in enabling adaptation to diverse environments.

### Identification of antimicrobial resistance genes (ARG)

Whole genome sequencing revealed the presence of multiple antibiotic resistance genes in all three *C. kerstersii* strains using ResFinder (Table 3). The tet(A) gene, which potentially conferred resistance by exporting tetracycline out of the cell in exchange for protons, was identified [44]. The strains possessed mph(E), mph(F), and msr (E) genes. The mph(E) and mph(F) encode macrolide phosphotransferase enzymes that likely inactivate macrolides through phosphorylation [45]. The msr(E) gene may confer resistance to macrolides, lincosamides, and streptogramin B [46]. The presence of the aph(3’’)-Ib and aph(6)-Id genes suggests resistance to streptomycin. These genes are often linked and encode enzymes that inactivate streptomycin [47]. The presence of sul2 gene confer resistance to sulfonamides.

**Table 3.**
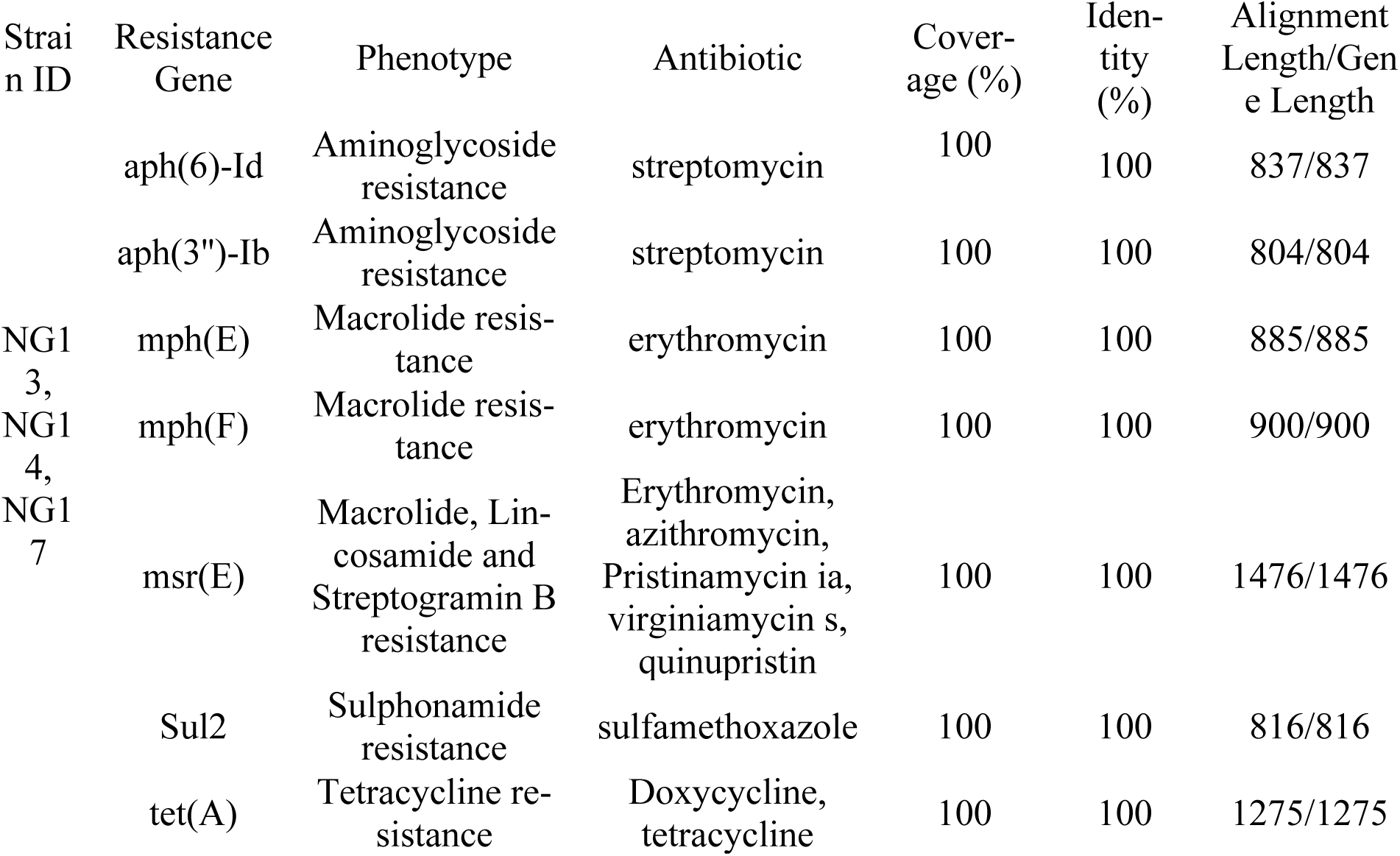
Antimicrobial resistance genes associated with the three strains of *C. kerstersii*.

We found macA, macB and tolC encoding for macrolide export protein MacA, macrolide export ATP-binding/permease protein MacB and outer membrane protein TolC, respectively. This MacAB-TolC comprise tripartite efflux system which is described to contribute in resistance to macrolide class of antibiotics in E. coli and other Gram-negative pathogenic bacteria [48]. This complex is also responsible for transportation of, protoporphyrin [49] and lipopolysaccharides [50] and virulence factors i.e. enterotoxin II [51]. Besides these, a number of multidrug resistance protein encoding genes are also present in the genomes of *C. kerstersii* strains (Supplementary Table 1).

### Adaptive Mechanism and Virulence

*C. kerstersii* can adapt to diverse environments, and annotating the genomes revealed some of the genes that enable them to withstand environmental stressors, including polyphosphate kinase (ppk). Polyphosphate (polyP) has previously been linked to osmotic and nutritional stress response in *E. coli* and bacterial growth, survival, and pathogenicity in other bacteria [52,53]. The presence of this enzyme explains how *C. kerstersii* is equipped to withstand osmotic stresses in various aquatic environments like blood, urine, or peritoneal fluid. Polyphosphate reserves in *Vibrio cholerae* improve the bacterium’s resistance to environmental stressors, especially when there is a shortage of phosphate (Jahid et al. 2006). The fact that *C. kerstersii* has polyphosphate kinase indicates that how it is adapted to survive in low-phosphate conditions, like those found in excrement in this study. PolyP has also been reported to be related with virulence including motility and biofilm formation in few studies [54–56].

The *C. kerstersii* strains of the study possess *osmY*, a periplasmic protein which is strongly induced by hyperosmotic stimuli. like *E.coli*, *C. kerstersii* may withstand external osmotic pressure like in bladder or colon and Yim et al. reported that *E. coli* express osmY proteins in such conditions [57]. The outer membrane protein A (*ompA*) precursor of *C. kerstersii* may enable it to withstand high osmolarity as the protein equips *E. coli* to survive in acidic environment, high osmolarity, and pooled human serum [58]. *OmpA* has also been associated with virulence factors like adhesion, invasion, intracellular survival, host defense evasion, and these activities are especially important in the urogenital, respiratory, or nervous system diseases [59]. The inner membrane protein required for strong colonization of the gut is encoded by the *cvpA* gene [60], and the presence of this gene in *C. kerstersii* strains of the study explains its adaptability in gut of the diarrheal patients. Additionally, a colicin V producing gene (*cvpA*), crucial for gut colonization, is present in *C. kerstersii* strains [60,61] and the presence of this gene in *C. kerstersii* strains of the study explains its adaptability in gut of the diarrheal patients.

Analysis using the Virulence Factor Database (VFDB) identified key proteins potentially associated with pathogenicity in all three *C. kerstersii* strains (Table 4). Notable findings include proteins involved with type IV pili (T4P), type II secretion systems (T2SS), type VI secretion system (T6SS), general secretion pathways, and flagella. Notably, both NG13 and NG14 strains encoded proteins involved in T4P biogenesis, including PilB and PilF. Strain NG17 possessed a PilJ protein. T4P play a crucial role in pilus formation, adhesion, and bacterial motility, all of which contribute to pathogenesis. T4P play an important role in pilus formation, adhesion, and dynamics [62].

**Table 4.** Virulence profile of *C. kerstersii* strains.

| Strain ID | Virulence Factors | Length | E-value | Overall Identity |
| --- | --- | --- | --- | --- |
| NG13 | VFG000112 (pilB) type 4 fimbrial biogenesis protein PilB [Type IV pili] [Pseudomonas aeruginosa PAO1] | 1701 | 2e-18 | 88.8% (166/187) |
|  | VFG000233 (pilF) type IV pilus assembly ATPase protein [Type IV pili] [Neisseria meningitidis MC58] | 1677 | 9e-17 | 80.9% (169/209) |
|  | VFG013777 (pilB) type II secretion system protein E:General secretory system II, protein E, N-terminal [Type IV pili biosynthesis] [Pseudomonas syringae pv. syringae B728a] | 1695 | 6e-18 | 85.4% (105/123) |
|  | VFG014984 (algW) AlgW protein [Alginate regulation] [Pseudomonas aeruginosa PAO1] | 1170 | 2e-18 | 83.6% (127/152) |
|  | VFG025878 (tsr) methyl-accepting chemotaxis protein [Flagella] [Burkholderia cenocepacia J2315] | 1959 | 9e-14 | 82.6% (119/144) |
|  | VFG030697 (sugC) ABC transporter, ATP-binding protein SugC [Trehalose-recycling ABC transporter] [Mycobacterium intracellulare MOTT-02] | 1179 | 1e-15 | 85.6% (95/111) |
|  | VFG038316 (tapD/pilD) prepilin peptidase [exe] [Aeromonas hydrophila subsp. hydrophila ATCC 7966] | 873 | 1e-15 | 87.4% (83/95) |
|  | VFG043040 (tar/cheM) methyl accepting chemotaxis protein II [peritrichous flagella] [Salmonella enterica subsp. enterica serovar Typhimurium str. LT2] | 1662 | 4e-19 | 85.6% (107/125) |
|  | VFG000182 (xcpR) general secretion pathway protein E [xcp secretion system] [Pseudomonas aeruginosa PAO1] | 1509 | 2e-09 | 85.9% (134/156) |
|  | VFG002048 (gspE) general secretion pathway protein E [T2SS] [Shigella dysenteriae Sd197] | 1494 | 6e-07 | 93.0% (40/43) |
|  | VFG000114 (xcpA/pilD) type 4 prepilin peptidase PilD [Type IV pili] [Pseudomonas aeruginosa PAO1] | 873 | 6e-07 | 90.2% (46/51) |
|  | VFG043209 (cheD) methyl-accepting chemotaxis protein CheD [peritrichous flagella] [Yersinia enterocolitica subsp. enterocolitica 8081] | 1674 | 9e-12 | 82.7% (191/231) |
| NGCRVN 14 | VFG036295 (pilF) type IV pilus assembly protein [Type IV pili] [Neisseria lac- tamica 020-06] | 1677 | 2e-21 | 81.8% (171/209) |
|  | VFG019728 (pilB) type 4 fimbrial biogenesis protein PilB [Type IV pili biosynthesis] [*Pseudomonas aeruginosa* LESB58] | 1701 | 9e-21 | 85.4% (229/268) |
|  | VFG026971 (sigA/rpoV) RNA polymerase sigma factor [Sigma A] [*Mycobacterium tuberculosis* CAS/NITR204] | 1134 | 9e-24 | 84.2% (139/165) |
|  | VFG038432 (tapB) type IV-A pilus assembly ATPase PilB [Tap type IV pili] [*Aeromonas hydrophila* ML09-119] | 1707 | 5e-19 | 88.2% (150/170) |
|  | VFG043040 (tar/cheM) methyl accepting chemotaxis protein II [peritrichous flagella] [*Salmonella enterica* subsp. enterica serovar Typhimurium str. LT2] | 1662 | 5e-19 | 85.6% (107/125) |
|  | VFG014984 (algW) AlgW protein [Alginate regulation] [*Pseudomonas aeruginosa* PAO1] | 1170 | 2e-18 | 83.6% (127/152) |
|  | VFG030705 (sugC) sugar-transport ATP-binding protein ABC transporter SugC [Trehalose-recycling ABC transporter] [*Mycobacterium liflandii* 128FXT] | 1188 | 3e-17 | 86.4% (95/110) |
|  | VFG043209 (cheD) methyl-accepting chemotaxis protein CheD [peritrichous flagella] [*Yersinia enterocolitica* subsp. enterocolitica 8081] | 1674 | 9e-12 | 82.7% (191/231) |
|  | VFG002529 (tsr) methyl-accepting chemotaxis protein I [Flagella] [*Burkholderia pseudomallei* K96243] | 2022 | 1e-10 | 81.6% (115/141) |
|  | VFG000182 (xcpR) general secretion pathway protein E [xcp secretion system] [*Pseudomonas aeruginosa* PAO1] | 1509 | 2e-09 | 85.9% (134/156) |
|  | VFG002048 (gspE) general secretion pathway protein E [T2SS] [*Shigella dysenteriae* Sd197] | 1494 | 6e-07 | 93.0% (40/43) |
|  | VFG000114 (xcpA/pilD) type 4 prepilin peptidase PilD [Type IV pili] [*Pseudomonas aeruginosa* PAO1] | 873 | 6e-07 | 90.2% (46/51) |
| NGCRVN 17 | VFG002076 (clpV1) type VI secretion system AAA+ family ATPase [HSI-I] [*Pseudomonas aeruginosa* PAO1] | 2709 | 8e-08 | 86.8% (59/68) |
|  | VFG043092 (tar/cheM) methyl-accepting chemotaxis protein II [peritrichous flagella] [*Escherichia coli* O157:H7 str. EDL933] | 1662 | 3e-22 | 88.2% (97/110) |
|  | VFG021055 (mgtB) Magnesium transport ATPase, P-type 2 [Mg<sup>2+</sup> transport] [Salmonella enterica subsp. enterica serovar Enteritidis str. P125109] | 2727 | 5e-21 | 79.8% (233/292) |
|  | VFG014045 (pilJ) type IV pilus biogenesis protein PilJ [Type IV pili twitching motility related proteins] [Pseudomonas syringae pv. tomato str. DC3000] | 2043 | 2e-17 | 87.3% (89/102) |
|  | VFG025879 (tsr) methyl-accepting chemotaxis sensory transducer [Flagella] [Burkholderia cenocepacia MC0-3] | 1974 | 7e-14 | 82.6% (119/144) |
|  | VFG046110 (ECVR50_2114) putative inner membrane ABC-transporter [Yersiniabactin siderophore] [Escherichia coli VR50] | 1803 | 3e-13 | 84.0% (100/119) |
|  | VFG034062 (ybtP) ABC transporter ATP-binding/permease rprotein [Yersiniabactin siderophore] [Escherichia coli O44:H18 042] | 1803 | 3e-13 | 84.0% (100/119) |
|  | VFG001231 (chpA) still frameshift probable component of chemotactic signal transduction system [Type IV pili] [Pseudomonas aeruginosa PAO1] | 7419 | 8e-08 | 95.0% (38/40) |

The presence of genes encoding proteins involved in flagellar morphogenesis, assembly, and chemotaxis (Supplementary Table 1), emphasize the role of mobility in bacterial pathogenic strategies. Bacteria navigate their environments using a combination of motility and chemotaxis. Motility allows them to actively seek out nutrients, while chemotaxis acts like a “chemical GPS,” guiding them toward favorable conditions and allows bacteria to rapidly adapt to changing environmental conditions [63]. In pathogenic bacteria, chemotaxis contributes to host invasion and infection by allowing the bacteria to sense and move towards host tissues and nutrients [64].

In general, there are noticeably few classical virulence determinants revealed by VFDB analysis. A thorough examination of the *C. kerstersii* genome, however, revealed a number of toxin coding sequences (CDS), including zona occludens toxin (zot) protein-encoding sequences that may play virulence-related activities. Another example is apxIB, which encodes Toxin RTX-I translocation ATP-binding protein, involved in the transport of the toxin RTX-I as well as that of RTX-II. RTX (Repeats-in-toxin) toxins are glycine– and aspartate-rich repeats containing exoproteins secreted by type I secretion system of Gram-negative bacteria[65]. One classical RTX protein is hemolysin which is encoded by tlyA in the genomes of *C. kerstersii* strains. The members of the RTX toxin family are important virulence factors of many Gram-negative bacteria [65]. Another gene is ratA which encodes Ribosome association toxin RatA. This protein is the toxic component of type II toxin-antitoxin (TA) system. In *E.coli* association of RatA protein with 50S subunit of ribosome prevents the binding of small subunit 30S, thereby preventing the formation of 70S ribosomes leading to decrease in protein translation [66].

The *C. kerstersii* strains possess genes that encode proteins involved in alginate biosynthesis pathway, Phosphomannomutase/phosphoglucomutase, Poly(beta-D-mannuronate) C5 epimerase 1, Poly(beta-D-mannuronate) C5 epimerase 5 encoded by algC, algE, algE1, respectively. A negative regulator of alginate synthesis called alginate biosynthesis sensor protein Kin**B**, encoding gene kinB is also present in the genome. AlgC protein plays a vital role in producing alginate, LPS and rhamnolipid. three important *P. aeruginosa* virulence-associated factors Alginate producing *P. aeruginosa* strains have a mucoid phenotype and are protected from antibiotics and other host defense components [67].

Clp protease (caseinolytic protease; ClpP) is a highly conserved serine protease found in many bacteria. This ATP-dependent protease functions as a two-component complex consisting of a regulatory ATPase subunit and a proteolytic subunit. While the proteolytic subunit possesses some intrinsic catalytic activity, both components are essential for optimal enzymatic function in the presence of ATP. We identified genes encoding various components of the Clp protease complex, including clpP and clpP1 (putative Clp proteolytic core), clpA, clpC, and clpX (Clp-ATPases), and clpS (adaptor protein). Clp proteases play crucial roles in various cellular processes, including the degradation of misfolded proteins, regulation of short-lived proteins, and housekeeping removal of dysfunctional proteins. They also control cell growth and target DNA-binding proteins from starved cells [68,69]. ClpP has also been linked to the tight regulation of virulence genes in both Gram positive and Gram negative pathogens such as Staphylococcus aureus, Listeria monocytogenes and Salmonella typhimurium. Therefore, the presence of Clp protease complex encoding genes indicate the contribution of this system in the pathogenic lifestyle of the *C. kerstersii* strains [69,70].

Iron is often limited in host environments as part of the innate immune response, so bacteria need to actively scavenge iron from sources like hemoglobin. This iron homeostasis is closely linked to bacterial virulence, stress resistance, and overall fitness. We examined the *C. kerstersii* genomes and found that a number of genes are present to encode enzymes and proteins involved in iron uptake mechanisms. Enterobactin is a siderophore that can extract iron from host iron-binding proteins like transferrin. Haemoglobin-haptoglobin binding proteins allow bacteria to retrieve heme iron from haemoglobin, a major iron source in the host [71]. Iron acquisition regulators like Fur control the expression of iron uptake systems in response to iron availability, allowing bacteria to tightly regulate iron uptake to avoid toxicity while ensuring sufficient iron for growth [71] All these proteins are key adaptations that enable *C. kerstersii* to efficiently scavenge iron from the host, a critical nutrient for growth and virulence. In addition, several genes encode key enzymes in the heme biosynthesis pathway that is essential for bacterial growth, metabolism, and virulence. Targeting these enzymes could be a potential antimicrobial strategy.

A cluster of genes involved in copper homeostasis was found in the annotated genomes of *C. kerstersii* strains (Table 5). Li et al. mentioned that a crucial factor in the pathophysiology of bacteria is copper homeostasis, which involves the complex interaction of bacterial survival strategies and host defenses [72]. To fight invasive pathogens, host macrophages deliberately create a copper overdose environment, exposing a host-driven defense mechanism (2019). Copper homeostasis-related genes, such as copA (efflux system), copZ (copper chaperone), cueO (copper oxidase), copS (sensor protein), and copR (transcriptional activator), were present in all three of the *C. kerstersii* strains included in this study. Notably, studies have shown that loss of the copper efflux pathway (CopA) in certain bacteria, such as S. pneumoniae, reduces virulence during infection [73]. This discovery implies that the pathogenic potential of *C. kerstersii* may depend on its capacity to maintain copper homeostasis.

**Table 5.**
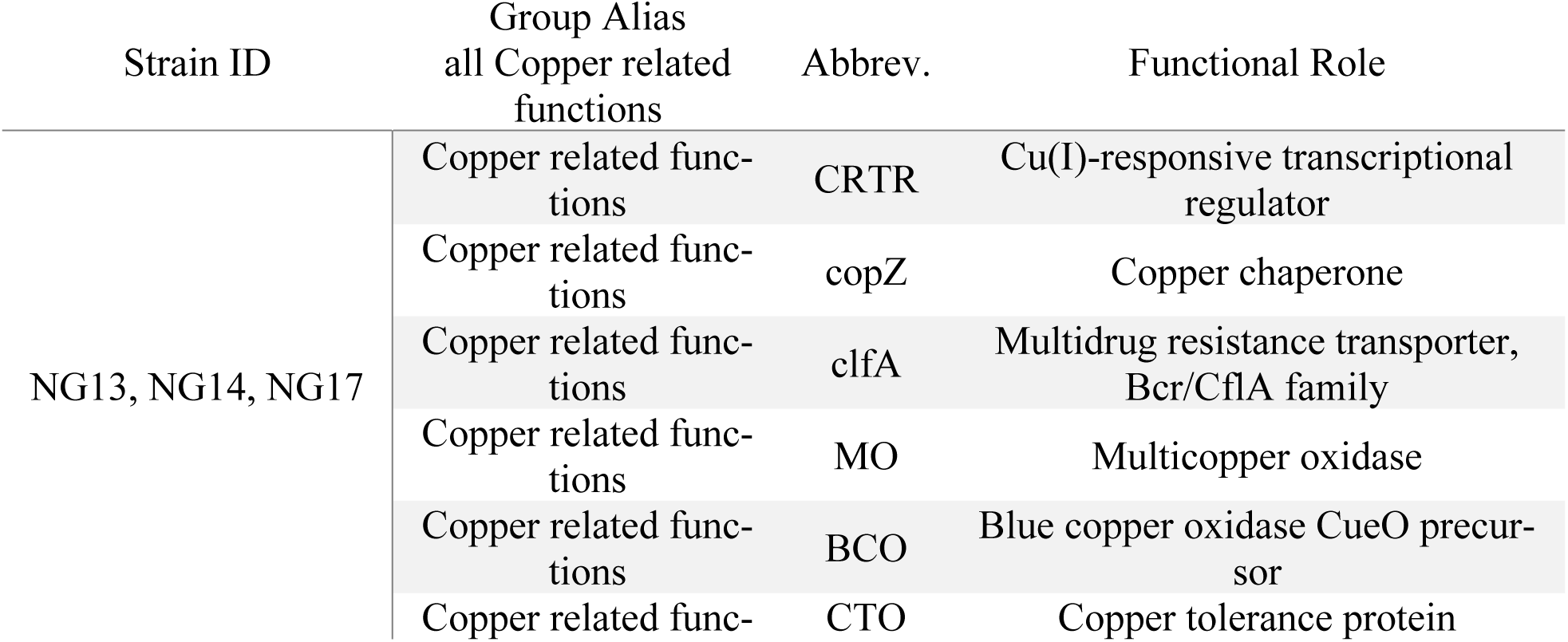
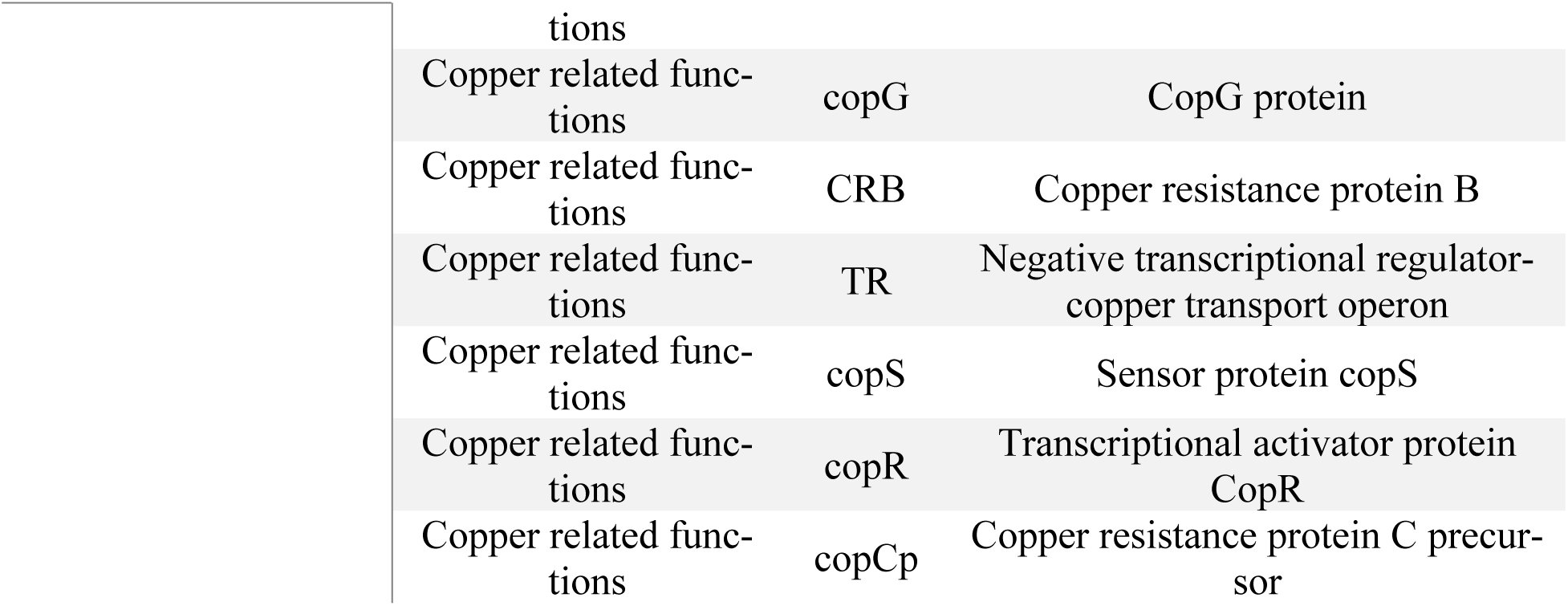
Genes related to copper homeostasis in *C. kerstersii* strains.

### Secretion systems

Protein export systems are responsible for regulating the secretion of different types of proteins across the cell envelope and thus has an important role in the colonization of bacteria living in various niches. *C. kerstersii* genomes were examined for the protein export and secretion system genes. In *Comamonas* species, the Sec (secretion) and Tat (twin-arginine translocation) pathways are essential for protein transport across the cytoplasmic membrane [74]. We found SecA, SecB, SecYEG encoding genes, although these genes are not present in a cluster. In contrast, tatABC genes encoding the components of the Tat apparatus, are present in a cluster (Supplementary table 1). The Sec pathway typically handles the translocation of unfolded proteins, whereas the Tat pathway is specialized for transporting fully folded proteins. Most proteins transported by the Sec and Tat pathways remain within the periplasm or inner membrane and may be transported in the extracellular environment through type II or type V secretion systems [75]

The genes epsE, gspF and outD encode components of type II secretion system and genes to encode Chaperone-Usher pathway (supplementary table 1). The chaperone–usher (CU) pathway is the most widespread method for assembling adhesive surface pili on the surface of Gramnegative bacteria. For uropathogenic *Escherichia coli* and *Pseudomonas aeruginosa,* these adhesive pili mediated localization and host colonization and biofilm formation, are considered as important virulence factors [75,76].

### Plasmids and Prophages

In each of the three strains used in this investigation, PlasmidFinder located a 157bp long fragment identical to plasmids of IncQ family. 100% identity was found in the region between 5741 and 6536 base pairs (bp) in the plasmid when it was compared using BLAST against the E. coli plasmid RSF1010 (Accession no. M28829). The distinct strand-displacement replication mechanism of the IncQ-family of plasmids allows them to work in a broad range of bacterial hosts and their high mobilization potential enables these plasmids to exhibit great promiscuity [77]. Several studies have mentioned plasmid IncQ to be multidrug resistant carrying *tetA*, Sul or carbapenem resistance gene blaKPC-2 [78,79] although the IncQ1 plasmid of the studied strain do not contain any resistance genes.

PHASTEST and Phigaro analysis identified prophage regions in all three *C. kerstersii* strains (Figures 3 and 4). Each prophage region was approximately 63.6 kb in size with a GC content of ∼58% and showed similarity to Pseudomonas phage F116 of the Podoviridae family. These regions encoded proteins involved in phage replication (integrase, repressor, terminases), regulation (regulatory protein), and lysis (lysis proteins, tail proteins, holin) and contained a significant number of hypothetical proteins.

**Fig. 3.**
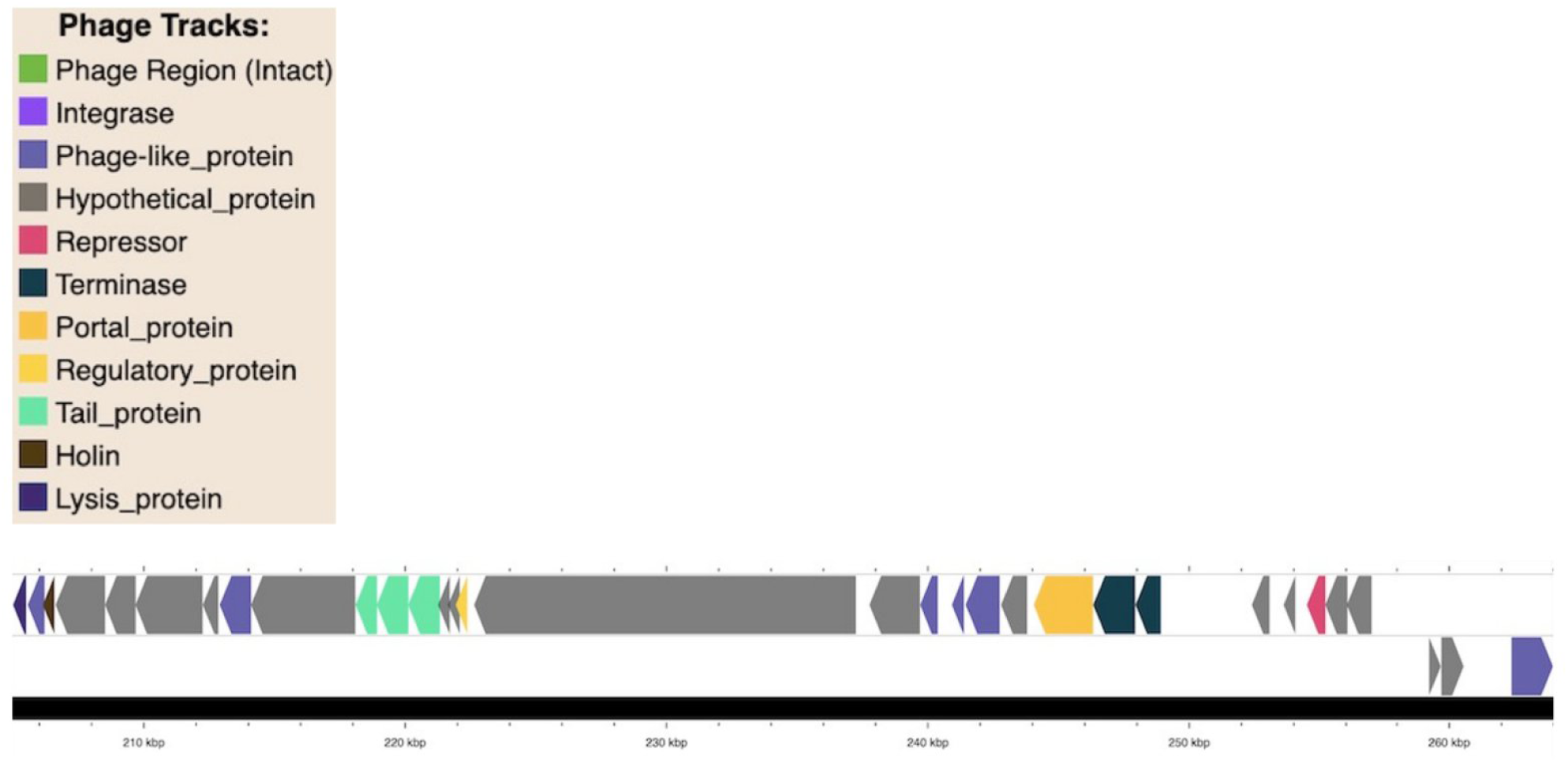
An intact phage region resembling *Pseudomonas* phage F116, identified using PHASTEST.

**Fig. 4.**
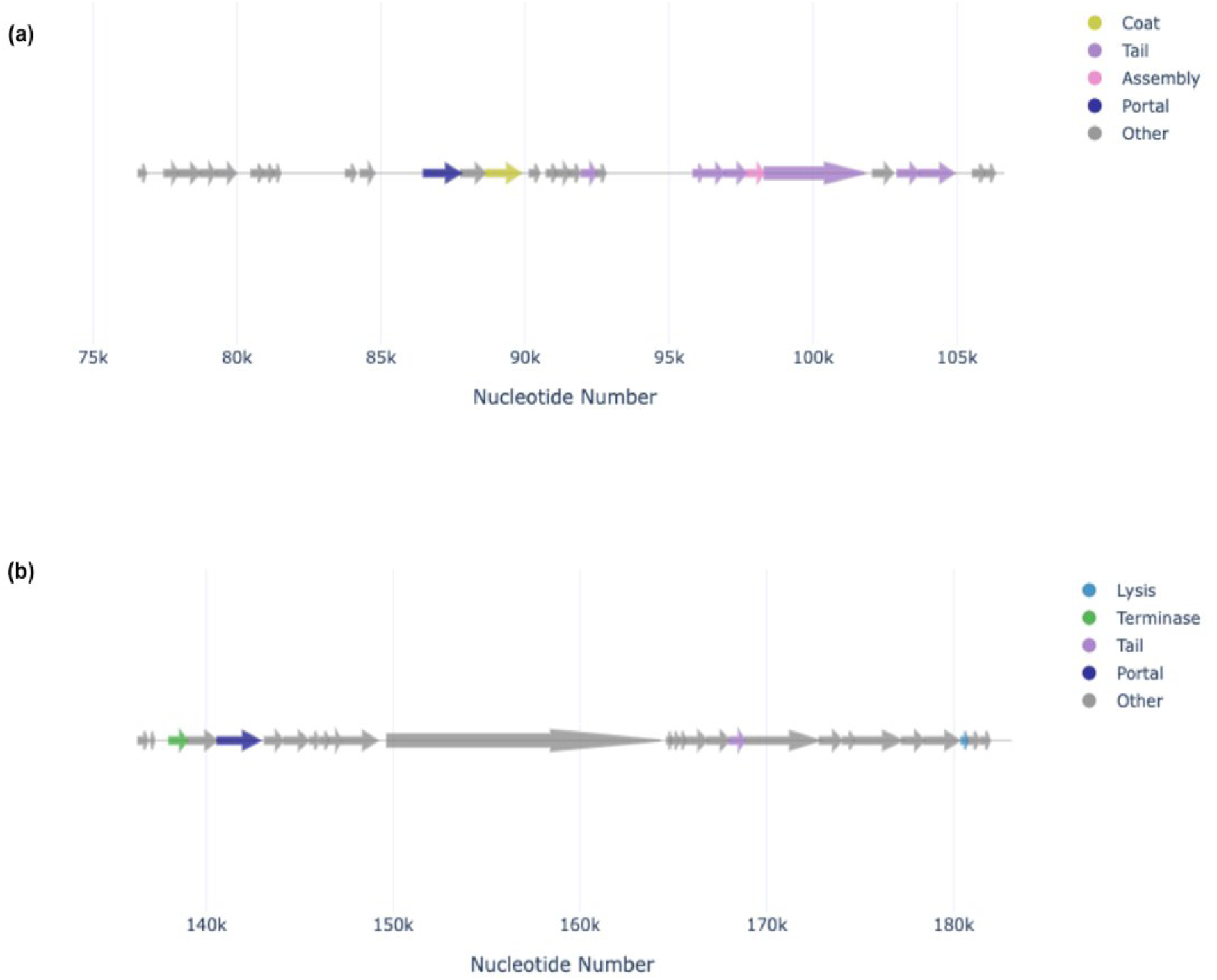
A phage region belonging to (a) Siphoviridae family and (b) Podoviridae family.

Interestingly, Phigaro identified an additional prophage region belonging to the Siphoviridae family alongside the Podoviridae region in each strain (Figure 4a). This region encoded phage assembly proteins (tail, portal, coat proteins). Multiple prophage regions and a significant number of hypothetical proteins within them suggest potential roles in *C. kerstersii* fitness. Further investigation of these hypothetical proteins may provide valuable insights into their specific functions.

### *Comamonas kerstersii* Host-Pathogen PPI Network

Following the BLASTp alignment of clustered *Comamonas* protein sequences with bacterial proteins from the PHISTO database, 1,269 homologous *Comamonas* proteins were identified. These proteins corresponded to known human–bacterial interactions in PHISTO, leading to the prediction of 2,119 protein–protein interactions (PPIs) between *Comamonas* and human proteins. The PPI network analysis identified the top 10 *Comamonas* hub proteins based on degree centrality. These proteins were Q5NF74, Q7CGS9, Q7CHZ0, Q5NGF1, Q5NFP9, Q8CZZ7, Q8D0D4, Q5NIP6, Q7CKZ7, and Q5NIP5. Together, these hub proteins interacted with 172 unique human proteins. This formed a subnetwork consisting of 184 predicted *Comamonas*–human protein–protein interactions.

## Discussion

*C. kerstersii is* becoming a prominent clinical and environmental pathogen due to its broad illness spectrum, ubiquity, adaptability in various environments and resistance capabilities. These characteristics, which indicate that *C. kerstersii* is highly diverse, call for a deeper comprehension of how the species has adapted to fill various niches. *C. kerstersii* came to light in Bangladesh for the first time; thus, genomic analysis and understanding the behavior of this bacterium is crucial for mitigating potential public health risks associated with its infections.

Phylogenetic tree construction with the other available nine strains of *C. kerstersii* reveals its clonal nature, diversity, and adaptability to different niches. Clustering of NG13, NG14, and NG17 together demonstrate their clonal nature, explaining the identical genotypical features. Several studies have mentioned the phylogeny of *C. kerstersii*, for instance Wu et al. (2016), Zhang et al. (2023) and Jiang et al. (2018) constructed phylogenetic tree, however, the construction of these phylogenies was between the species of the genus *Comamonas* and not specifically between different strains of *C. kerstersii* [17, 24, 80]. The limited number of samples of *C. kerstersii* is one of the notable limitations of this study and as well as other investigations to fully understand the true nature and genetic diversity of this species. To understand the genetic diversity of this species at spatial-temporal level and to obtain a more accurate picture of the genetic variety of *C. kerstersii* circulating across the country by significantly increasing the number of samples from various sources and places.

The strains of the study have been isolated from clinical samples. We have identified several virulence factors other than T4P and T6SS. It is plausible that these strains have acquired the virulent characteristics from the other species that may have coexisted in the infection milieu. The toxin protein zot is responsible for diarrhea or the holin protein present in the phage region can also induce the pathogenicity. The enhanced release of extracellular toxins in *E. coli*, including *SheA* and *Stx1*, has been linked to phage-encoded holins [81]. According to a number of investigations, holins most likely also have a significant function in the production of biofilms [82]. For instance, it is known that the *Staphylococcus aureus cid* and *lrg* operons regulate cell lysis and the release of genomic DNA, which eventually becomes a structural element of the biofilm matrix and are thus implicated in the production of biofilms [82]. Alternatively, they may aid in the infection process through systems not generally linked to virulence but through resistance mechanisms like regulation of copper homeostasis.

As a relatively new opportunistic pathogen, *C. kerstersii* has received little attention in scientific research. Only two papers have explored and reported the genomic analysis of this species thus far suggesting that the genomic microbial databases might have incomplete information on *C. kerstersii.* This lack of information may have made it difficult to identify every gene connected to the organism. The presence of many hypothetical proteins has been discovered by the annotated data of the studied strains. These proteins are hypothetical gene products, the roles of which are neither entirely understood nor confirmed by experiments. Even though they are speculative, these potential proteins can provide important information about the genetic composition of *C. kerstersii* and could be useful research targets to clarify their functions in pathogenicity and antibiotic resistance.

## Conclusion

Most of the researchers focus on well-known pathogens like *V. cholerae* and *E. coli* frequently when isolating microorganisms from diarrheal patients. However, recent advances in high throughput genomics facilities have identified other potential pathogens from diarrheal samples. It is imperative that we adapt the state-of-the-art genomics technologies and expand our research focus to consider other possible pathogens like *C. kerstersii.* Significant insights into the role of *C. kerstersii* in diarrheal disorders can be gained from an understanding of its pathophysiology and genetic features. By uncovering previously unknown infections, this widened focus may contribute to the development of more thorough diagnostic procedures and focused therapeutic methods. Researchers can improve public health responses and our understanding of diarrheal disorders by investigating the genomes of *C. kerstersii* to identify its virulence factors, resistance mechanisms, and potential epidemiological consequences.

## Ethical Approval

All isolates analyzed in this study were obtained for diagnostic purposes from a local tertiary hospital. All experiments and methodologies were conducted in compliance with relevant guidelines and regulations. The North South University (NSU) Institutional Review Board (IRB) / Ethical Review Committee (ERC) approved all experimental protocols, under protocol No. 2020/OR-NSU/IRB/0701

## Data Availibility

The 3 *Comamonas kerstersii* genome sequences and analysis from this study have been submitted to GenBank database, with accession numbers: JBEBZE000000000 (NG17), JBE-BZF000000000 (NG13), and JBEBZG000000000(NG14).

## Authors’ Contribution

**Study Design:** NIR, FS, FK, SAH, AS, TSH, MH

**Sample Collection:** FS, JA, SNT, AR

**Experimental Procedures:** NIR, FS, FK, SAH, PD, AH

**Data Analysis and Interpretation:** NIR, FK, SS, SAH, AS, AH, MH

**Manuscript Drafting and Revision:** NIR, FS, FK, SS, AS, SAH, MH

**Supervision:** MH

**Corresponding Author:** MH

## Data Availability

All data produced in the present work are contained in the manuscript

